# Confounding factors affecting analysis of germline structural variants in pediatric solid tumors

**DOI:** 10.1101/2025.07.16.25331652

**Authors:** Pandurang Kolekar, Rebecca S. Kaufman, Nadezhda V. Terekhanova, Jinghui Zhang, John M. Maris, Sharon J. Diskin, Xiaotu Ma

## Abstract

We provide data showing the germline predisposition structural variants (SV) involving *MYCN* and *RAF1::TMEM40* reported by Gillani et al. (2025) in pediatric solid tumors are confounded by circulating tumor DNA (ctDNA) and enrichment for Hispanic or Latino ancestry, respectively. We suggest that future germline studies should ensure analyses of tumor-in-normal contamination for non-polymorphic markers and careful examination of population stratification for polymorphic markers to ensure clinical relevance.

## Main text

Although most childhood cancers are driven by somatic alterations (*1, 2*), pathogenic and likely pathogenic alterations can be identified in the non-cancerous tissues of childhood cancer patients reflecting *bonafide* germline inheritance or post-zygotic mosaicism (*3*). Identification of such germline events are of pivotal importance for disease surveillance, management, and genetic counseling. As such, there has been tremendous interest in the analysis of pathogenic germline alterations, such as the recent report by Gillani et al. (*4*). However, their results are confounded by factors including 1) ctDNA in the *MYCN* case and 2) population stratification for cases with SV polymorphisms involving *RAF1::TMEM40*. We present below our re-analysis of these cases.

### Presence of MYCN copy gain in germline samples confounded by circulating tumor DNA

The two neuroblastoma cases (sourced from Gabriella Miller Kids First pediatric research program, GMKF) described in Fig. 5F of Gillani et al. with proposed germline or post-zygotic mosaic *MYCN* gain are of particular interest. Due to availability of tumor sample, we present re-examination of the *MYCN* amplicon in case PT_V1Q9W1NW in the paired tumor and the normal samples (table S1; Supplementary Materials and Methods). The tumor sample is estimated to have ∼44 copies of *MYCN* based on read-depth coverage (Fig. 1A). This gain is clearly present in the peripheral blood sample (∼1 copy gain) but absent in the paternal and maternal buccal samples. The conclusion of germline nature in Gillani et al. is largely drawn from here.

**Fig. 1.**
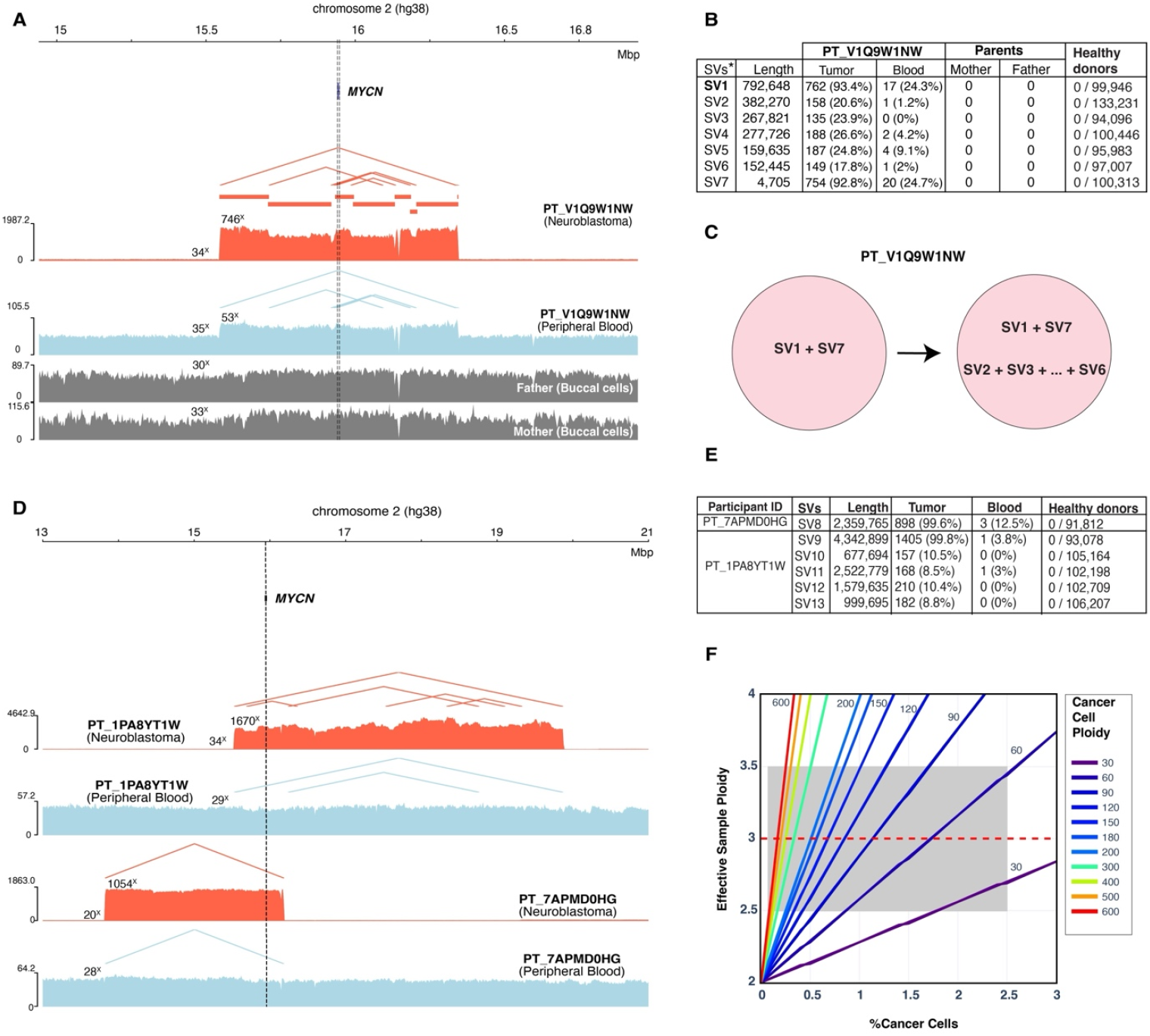
Circulating tumor DNA (ctDNA) confounds germline analysis of *MYCN* structural variation. (**A)** Whole genome sequencing (WGS) based read depth profiles of neuroblastoma (red) and normal sample (blue; peripheral blood; PB) for case PT_V1Q9W1NW along with parents’ normal samples (buccal cells; gray). Horizontal red lines indicate copy number events detected in the tumor sample, while red and blue triangle vertices indicate SVs events detected in tumor and normal samples, respectively. **(B)** Table showing SVs (shown are mutant read counts and mutant allele frequencies in parentheses) detected in tumor, matched normal, and parental buccal cells. SVs (SV1 to SV7) are listed in order of occurrence of their first breakpoint from left to right on chromosome 2 as shown in panel A. Also presented are genotyping results in WGS samples from 2,983 healthy donors. *: see actual SVs in table S2. **(C)** Schematic showing the proposed SV clonal evolution in the neuroblastoma tumor. **(D)** WGS read depth profiles of two additional cases from the same neuroblastoma cohort in Gillani et al. where ctDNA is detected from PB. (**E**) Table showing SV events detected in the two cases from (D). The definitions are the same as in the Fig. 1B. **(F)** Mathematical model showing relationship between cancer cell ploidy (CCP; here *MYCN* region), cancer cell fraction (x-axis; CCF), and effective sample ploidy (y-axis). The gray highlight indicates scenarios (low CCF and high CCP) where effective sample ploidy is ∼3. In panels A and D, vertical dashed lines indicate *MYCN* location, and numbers over coverage tracks denote median depth of corresponding regions.

Interestingly, the *MYCN* amplicon in the tumor sample appears to contain eight segments with varying magnitude of amplifications supported by concordant SV breakpoints (Figs. 1A and 1B). These included two clonal SVs (SV1 and SV7) and five subclonal SVs (SV2 to SV6) supported by ∼20% read counts of the clonal events (Figs. 1B and 1C; fig. S1; tables S2 and S3).

If the *MYCN* gain in the peripheral blood truly has germline origin, then the subclonal SVs in tumor should NOT be detectable in this specimen, a fact that we had used in a previous study to distinguish *bona fide* germline mosaicism from tumor-in-normal contamination (*3*). We therefore examined for the presence of these seven tumor SVs in the peripheral blood and were able to detect both the clonal and the subclonal SVs in peripheral blood DNA (tables S2 and S3). To rule out the possibility that sequencing errors may confound the observation, we searched against 2,983 WGS peripheral blood samples collected from survivors of childhood cancer and their matched community controls (*5*) and did not detect any SV reads out of cumulative ∼100,000X depth (Fig. 1B, tables S2 and S4). These analyses clearly indicated that the peripheral blood is contaminated with a low level of ctDNA from circulating tumor cells (CTC) or cell-free tumor DNA (cfDNA), a key biological premise for the liquid biopsy field. Indeed, neuroblastoma is known to have one of the highest ctDNA/CTC contents of all human cancers, and detection of somatic *MYCN* amplification in peripheral blood has been documented by many investigators (*6, 7*). Our mathematical modeling estimated the level of ctDNA to be ∼2%. Interestingly, we did not detect any additional clonal somatic single nucleotide variants (SNVs) and copy number variations (CNVs; this patient was diagnosed at age 2.8 years) in the tumor sample (*8*). The lack of additional clonal somatic markers is likely the reason for Gillani et al. to fail the tumor-in-normal contamination analysis (*3, 9*).

Using this above analysis, we detected ctDNA in two additional cases (PT_7APMD0HG and PT_1PA8YT1W) out of seven *MYCN* amplification cases with peripheral blood as the “normal germline” control DNA (Figs. 1D and 1E; tables S2, S5 and S6), totaling a detection rate of 43% (3/7). By contrast, we did not detect any (0% detection rate) ctDNA in buccal samples from the 39 *MYCN* cases (table S6). This is consistent with the assumption that ctDNA can be present in blood but not in buccal samples (*6, 10*).

To investigate when low ctDNA can be mistaken as germline events for high-copy-gain biomarkers, we have generated a mathematical chart where the local ploidy of the “germline” sample is modeled as a function of the actual cancer cell ploidy (CCP) and the cancer cell fraction (CCF), so that the effective sample ploidy would be ∼3 (Fig. 1F). Clearly, many scenarios with a high *MYCN* copy number combined with low ctDNA level can generate a local ploidy of 3, which in turn will lead to a false impression of “germline” status (gray highlight in Fig. 1F). Importantly, at these low ctDNA level of <3%, the current standard WGS at 30-60X coverage is not sufficiently powered for detecting somatic SNVs/Indels that can augment tumor-in-normal contamination analysis, resulting in mis-assignment of germline origin of a somatically acquired amplicon.

### Population stratification confounds RAF1-TMEM40 SV polymorphism analysis

We were also intrigued by the SV polymorphism involving duplication of *RAF1* and *TMEM40* described in Fig. 5C of Gillani et al. as being attributed to genetic predisposition to solid tumors based on its higher frequency in pediatric patients (3/1724; Fig. 2A, fig. S2, tables S7 and S8) versus controls (n=0 out of 4983; BioMe and MESA cohorts of NHLBI TOPMed described in Gillani et al). Interestingly, we discovered that the three index cases are enriched with Latino/admixed American ancestry inferred from global and local ancestry analyses of the *RAF1* and *TMEM40* locus on chr3p arm (Figs. 2A, 2B and 2C; table S9; Supplementary Materials and Methods). The proband case PT_ZT2NW6WA was particularly interesting as it appears to be European from global ancestry analysis but has Latino/admixed American ancestry at this SV locus upon local ancestry analysis (Figs. 2B and 2C; fig. S3A).

**Fig. 2.**
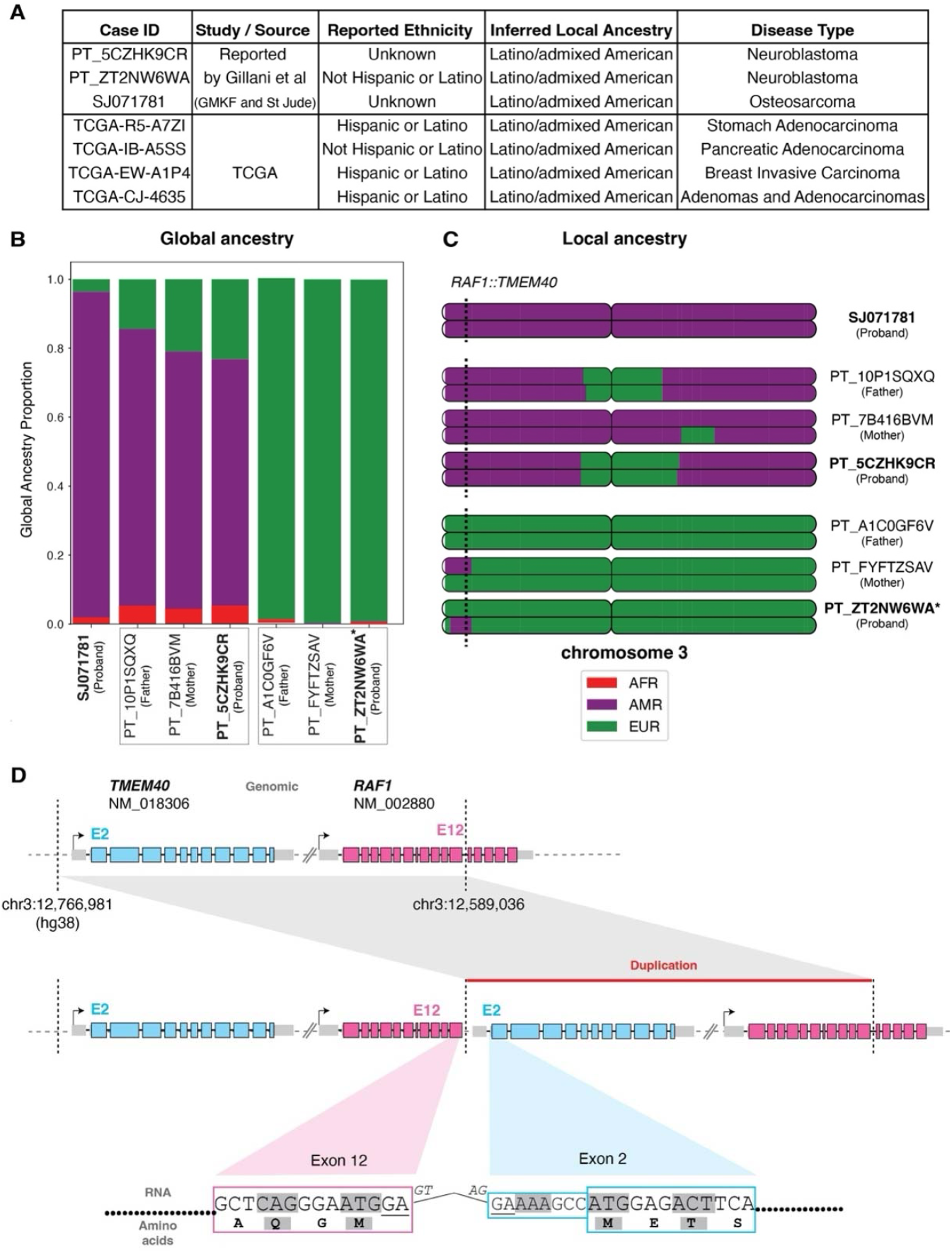
Population stratification confounds *RAF1::TMEM40* SV polymorphism analysis. (**A)**. In addition to the three cases reported by Gillani et al., we identified four TCGA cases carrying *RAF1::TMEM40* SV polymorphism. Self-reported ethnicity and genomically inferred local ancestry of *RAF1* and *TMEM40* locus in chr3p arm are listed along with cancer subtypes.Global and **(C)** Local ancestry analysis of probands carrying *RAF1::TMEM40* SV polymorphism. Cases marked in bold indicate probands. Related individuals from the same families are grouped in boxes shown along X-axis of global ancestry plot. Proband/parent status is also indicated under respective GMKF case IDs. Dashed vertical line across local ancestry plots indicates locus of *RAF1::TMEM40* on chromosome 3. Genetic ancestry abbreviations are shown in the combined color legend for both plots -AMR: Latino/admixed American (purple), AFR: African/African American (red), EUR: European (green). *: see fig. S3A for genome wide local ancestry analysis for case PT_ZT2NW6WA along with parents. (**D**) Pathogenicity analysis of *RAF1::TMEM40* SV. Shown are reference (top) and mutant (middle) genome of *RAF1*::*TMEM40* locus along with detailed gene structure of both genes and the 1-copy duplication region. Also shown is the exon junction (bottom) between *RAF1* (E12) and *TMEM40* (E2) induced by the SV polymorphism ascertained from 6 RNA-seq samples (see fig. S4 and table S10). The junction analysis indicated the fusion transcript is out-of-frame.

Owing to the relatively small sample size of their control cohort, we investigated the prevalence of this SV in populational SV database gnomAD (v4.1.0) and discovered a matching SV (ID: DUP_CHR3_76AC7D44; accessed May 10, 2025) with 42 alleles (n=126,078 alleles; 0.033%) (*11*). Interestingly, the carriers are also enriched with Latino/admixed American ancestry (74%; 31/42; Fisher’s test *P* = 10^−22^). Furthermore, this marker is >10 times more enriched in admixed Americans (0.2%; n= 12,586 alleles) than in other ancestry groups (<0.02%).

We next asked if individuals carrying this ultra-rare polymorphism can be found in adult cancer patients. We found four TCGA cases (n=9,447 WGS) diagnosed with different cancer subtypes (Fig. 2A, tables S7 and S8) harboring this SV polymorphism. Interestingly, all four cases were either self-reported to be from “Hispanic or Latino” ethnicity or inferred to have Latino/admixed American ancestry in *RAF1* and *TMEM40* locus on chr3p arm (case: TCGA-IB-A5SS, figs. S3B and S3C). These data further contradict the proposed predisposition to childhood solid tumors of this SV polymorphism by Gillani et al.

This SV is a tandem duplication predicted to generate *RAF1::TMEM40* fusion representing an out-of-frame transcript, exclusively between exon 12 of *RAF1* and exon 2 of *TMEM40* (Fig. 2D) (*12*), which is also detected in RNA-seq data of TCGA (table S10, figs. S4 and S5). Interestingly this fusion transcript is also found in pediatric leukemia patients (*13*). As such, the elevated expression level (Fig. 5D of Gillani et al.) of *RAF1* due to this SV is likely to be a byproduct of out-of-frame fusion transcript expression.

Together, our analyses highlight the importance of examining tumor-in-normal contamination and population stratification when analyzing candidates for germline cancer predisposition.

## Supplementary Materials and Methods

### Data for cases described in Gillani et al (2025) and supportive analyses

The participant IDs for the cases with germline or post-zygotic copy number gains (involving *MYCN* or *RAF1::TMEM40*) described in Fig. 5C and 5F of Gillani et al (*4*) were obtained from the authors (table S1). The WGS/RNA aligned reads in BAM format for proband (tumor and normal samples) or parents (normal samples) were downloaded from the Gabriella Miller Kids First (GMKF) project (https://kidsfirstdrc.org/) and St. Jude cloud (*14*) based on their respective participant IDs.

We also downloaded WGS BAM files for tumor-normal paired samples of 336 cases of the KF-NBL study (dbGaP study accession: phs001436.v1.p1) to detect additional cases demonstrating tumor-in-normal contamination by using SVs responsible for *MYCN* amplicon (table S5).

The WGS data from normal samples of 2,983 childhood cancer survivors sequenced as part of SJLIFE study (*5*) were downloaded from St. Jude cloud (https://www.stjude.cloud/) and used as control cohort to assess potential false positive SV genotyping (table S4).

The WGS/RNA data in BAM format from TCGA cohort were used to interrogate ethnicity bias of *RAF1::TMEM40* SV polymorphism. The “BAM Slicing Download” API available on Genomics Data Commons portal (https://portal.gdc.cancer.gov/) was used to download sliced portion of BAM files involving *RAF1::TMEM40* region padded with 500k bp on both side of the breakpoints.

### Bioinformatics and statistical analyses

The copy number and structural variants (SV) in WGS BAM files were detected using CONSERTING (*15*) and CREST (*16*) respectively. The SV breakpoints were manually confirmed using Integrative Genomics Viewer (IGV) v2.19.1 (*17*), followed by genotyping using SVInDelGenotyper (*18*). Only reads with >80% of bases having Phred score 30+ were used to quantify SV-supporting reads. The SNVs were detected using Bambino (*19*). Depth of coverage tracks were generated using pyGenomeTracks v3.5.1 (*20*). The samtools mpileup was used to compute the depth of coverage near SV breakpoints for diploid and *MYCN* amplicon regions (*21*). For case PT_V1Q9W1NW, the tumor sample is estimated to have ∼44 (i.e. 2*746x/34x) copies of *MYCN* whereas the normal sample appears to have three copies (i.e. one copy gain; 2*53x/35x).

The global and local ancestries for three cases carrying *RAF1::TMEM40* SV as described in Gillani et al (2025) were inferred as follows. The variants from 1000 genomes reference samples(*22*) were filtered with PLINK v1.9 (*23*) using --geno 0.01 --mind 0.01 --hardy --maf 0.01 -- biallelic-only strict and for LD using --indep-pairwise 50 5 0.5. These filtered SNPs were extracted from cases using PLINK v1.9. Genotypes were phased with Beagle v5.5 (*24*). Local ancestries were identified with RFMix v2.03-r0 (*25*) with default settings across the genome considering reference 1000 genome populations of EUR, AMR and AFR. The proportion of the genome mapped to specific ancestries was used to infer global ancestry. Local and global ancestries were then extracted from RFMix results and plotted with RFMIX2-Pipeline-to-plot (*26*).

The inferred ancestry and ethnicity information for the TCGA cases was obtained from The Cancer Genetic Ancestry Atlas (TCGAA) portal (http://fcgportal.org/TCGAA/) described in (*27*).

NeoVersioner (*28*) was used to assess the functional impact of *RAF1::TMEM40* fusion (resulting from the SV polymorphism) in RNA BAMs of available samples.

### Mathematical modelling of cancer cell fraction, cancer cell ploidy, and effective sample ploidy

The relationship between cancer cell fraction (CCF), cancer cell ploidy (CCP) and the effective sample ploidy can be modelled as follows:

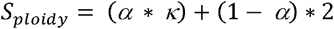

where S_ploidy_ denotes effective sample ploidy, α denotes cancer cell fraction (CCF), κ denotes cancer cell ploidy (CCP), and 1-α denotes normal cell fraction.

Because ctDNA level is typically low, CCF is generally low in normal controls.

Sample level B-allele frequency (BAF, where B denotes gained allele) can be modelled as follows:

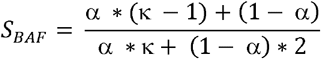

The above two equations are used to estimate the CCP in cancer cells using tumor sample that in turn was used to estimate the CCF in normal controls.

## Data Availability

All raw genome sequencing data analyzed in this study is described in Gillani et al (2025) and are hosted by dbGaP (https://dbgap.ncbi.nlm.nih.gov/; cohorts: MESA, BioMe, GMKF), St. Jude Cloud (https://www.stjude.cloud/), The Cancer Genome Atlas (TCGA; https://portal.gdc.cancer.gov/). Accession numbers for respective cohort and cases are provided in the supplementary materials and methods or tables S1, S4, S5 and S7.

https://www.stjude.cloud/

https://portal.gdc.cancer.gov/

https://kidsfirstdrc.org/)

## Acknowledgments

We acknowledge co-first and corresponding authors of Gillani et. al (2025) for kindly sharing the participant IDs for cases with *MYCN* amplification and tandem duplication involving *RAF1* and *TMEM40*.

## Author contributions

Conceptualization: PK, XM

Methodology: PK, RSK, NVT, SJD

Formal analysis: PK, RSK, NVT, JZ, SJD, JMM, XM

Investigation: PK, RSK, NVT, JZ, SJD, JMM, XM

Visualization: PK, RSK, NVT

Funding acquisition: XM, SJD

Project administration: XM, SJD, JMM

Supervision: XM, JZ, SJD, JMM

Writing – original draft: PK, XM

Writing – review & editing: PK, RSK, NVT, JZ, SJD, JMM, XM

## Competing interests

None

### Data and materials availability

All raw genome sequencing data analyzed in this study is described in Gillani et al (2025) and are hosted by dbGaP (https://dbgap.ncbi.nlm.nih.gov/; cohorts: MESA, BioMe, GMKF), St. Jude Cloud (https://www.stjude.cloud/), The Cancer Genome Atlas (TCGA; https://portal.gdc.cancer.gov/). Accession numbers for respective cohort and cases are provided in the supplementary materials and methods or tables S1, S4, S5 and S7. The software and databases used in this study are publicly available and cited appropriately in the main text or supplementary materials and methods.

