## Supplementary figures and images for "Confounding factors affecting analysis of germline structural variants in pediatric solid tumors"

### Fig. S1

A

**PT_V1Q9W1NW** (SV1-SV7)

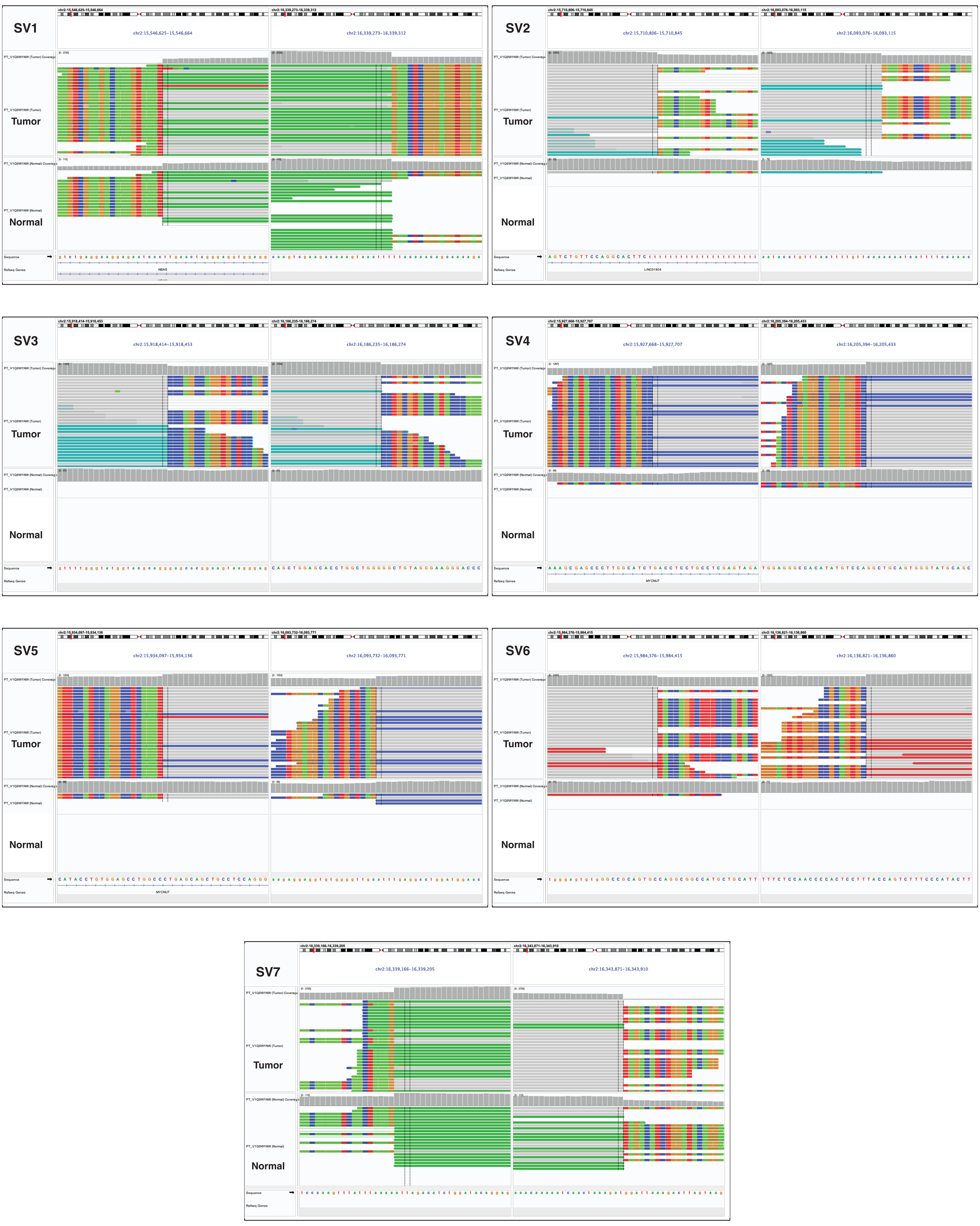


B

**PT_7APMD0HG** (SV8)

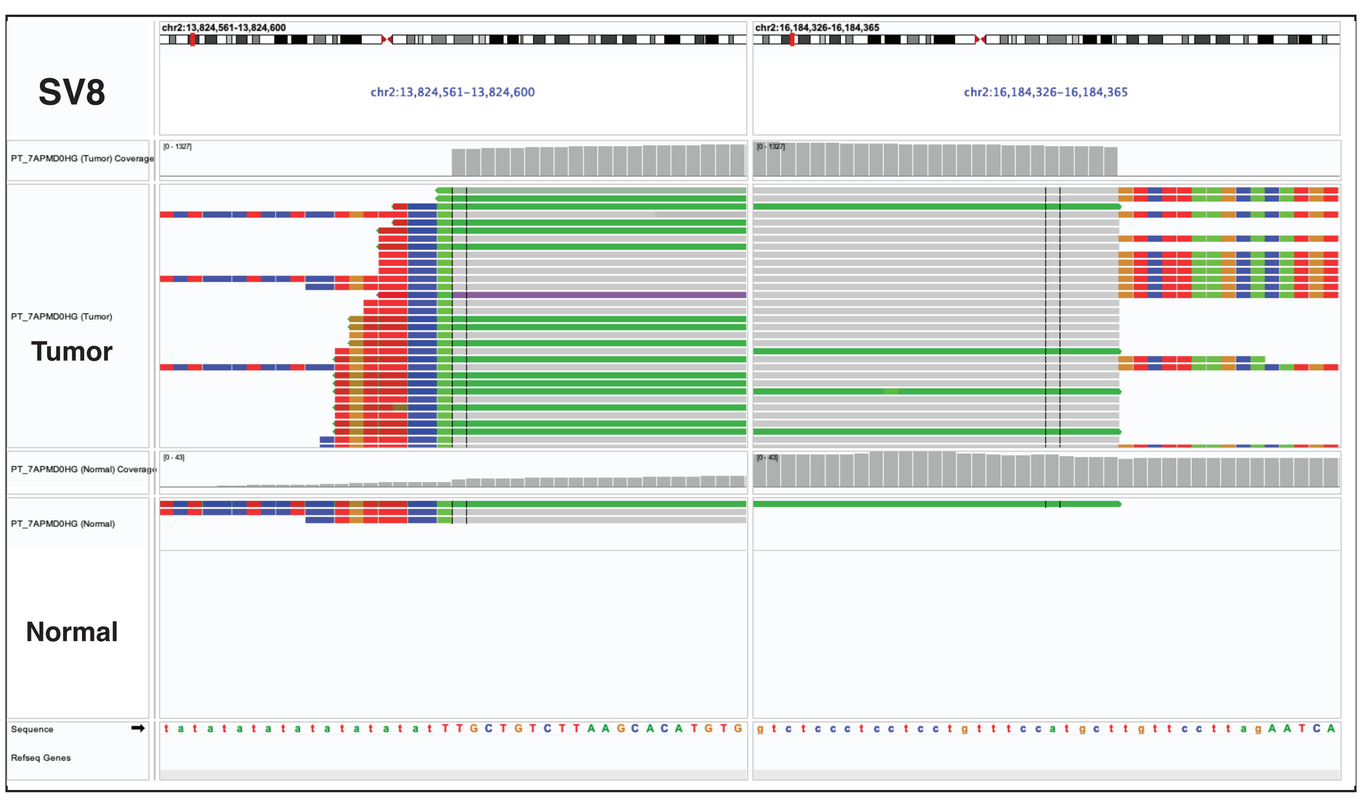


C

**PT_1PA8YT1W** (SV9-SV13)

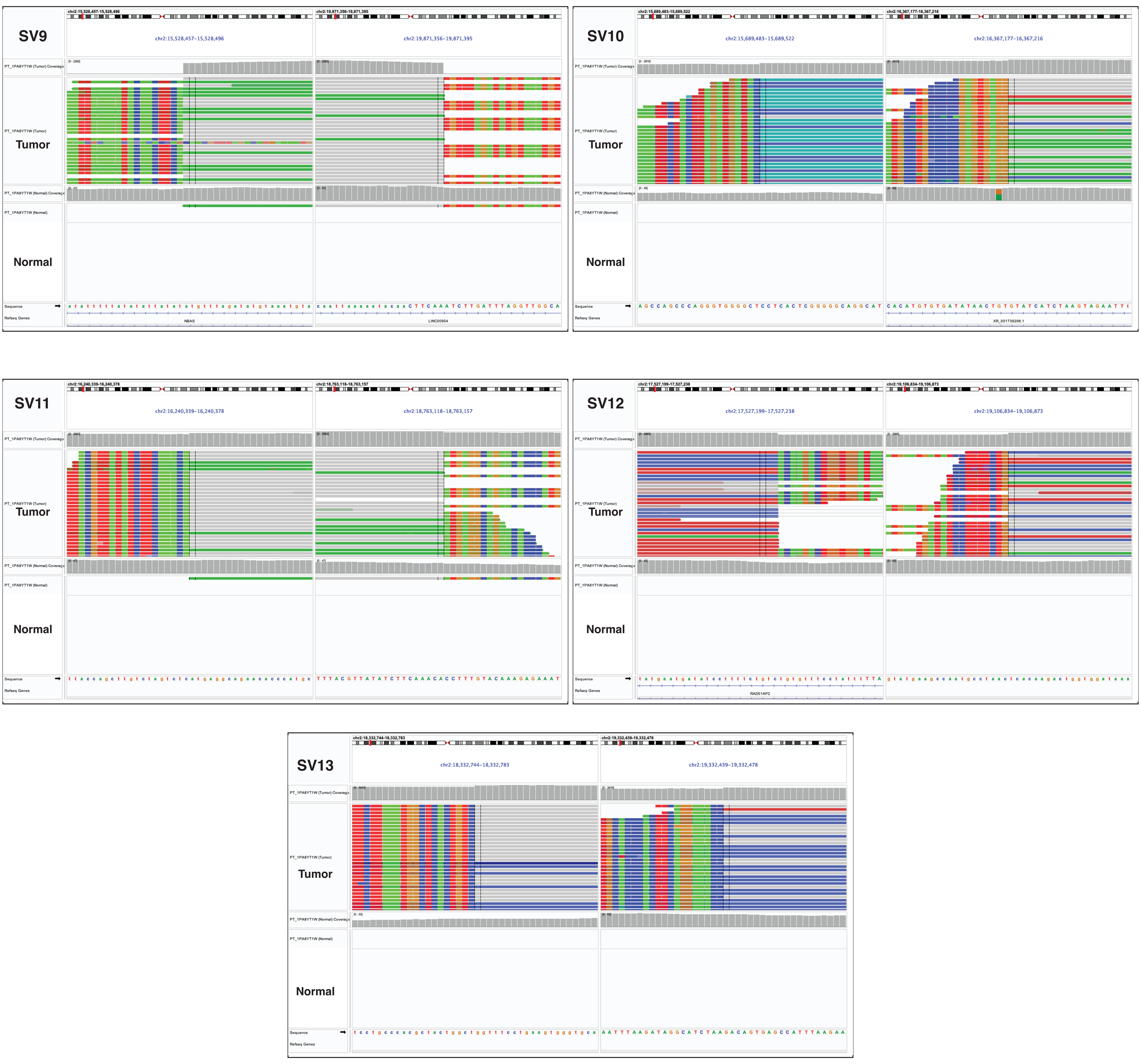

### Fig. S2

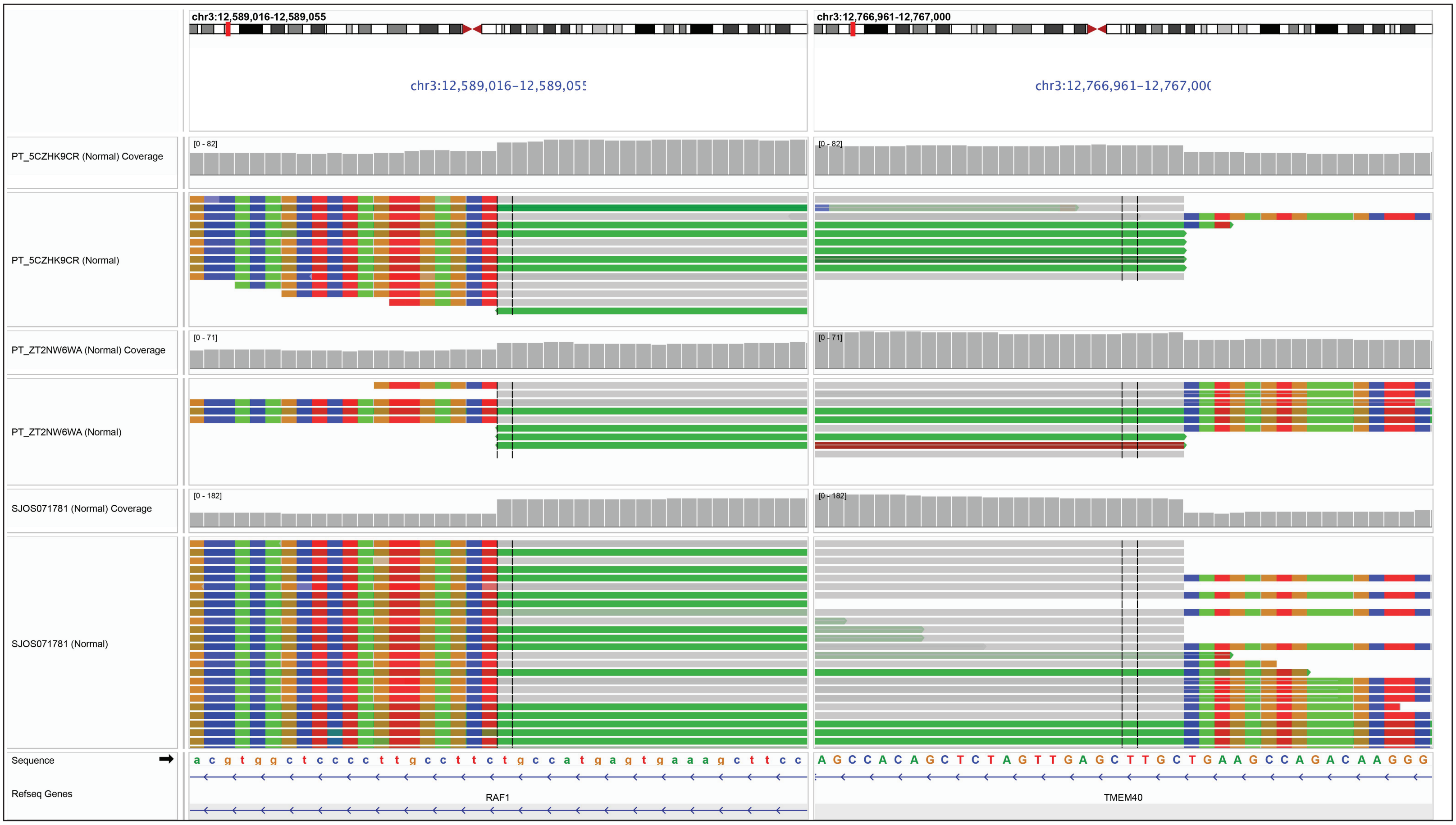

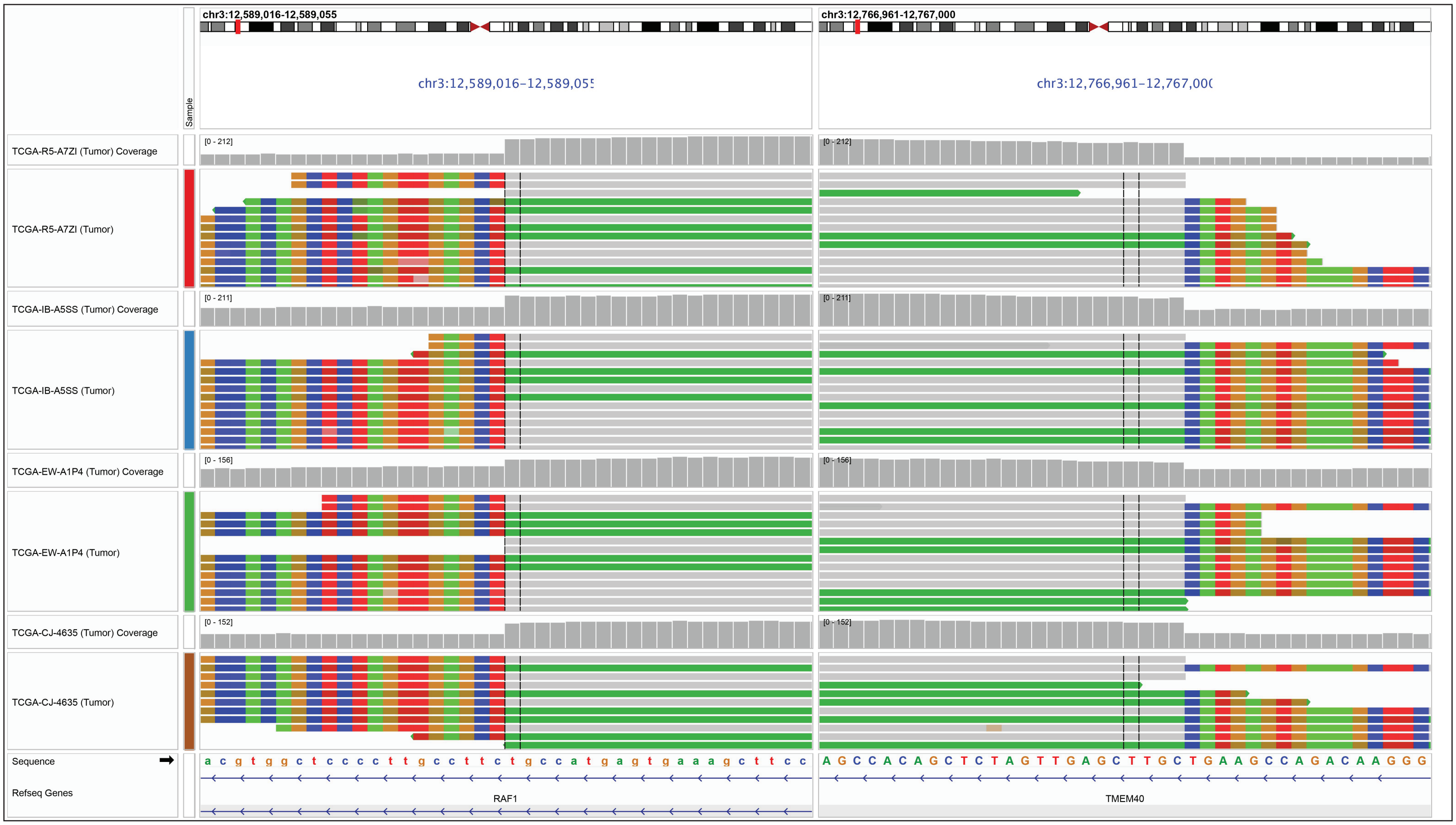

### Fig. S3

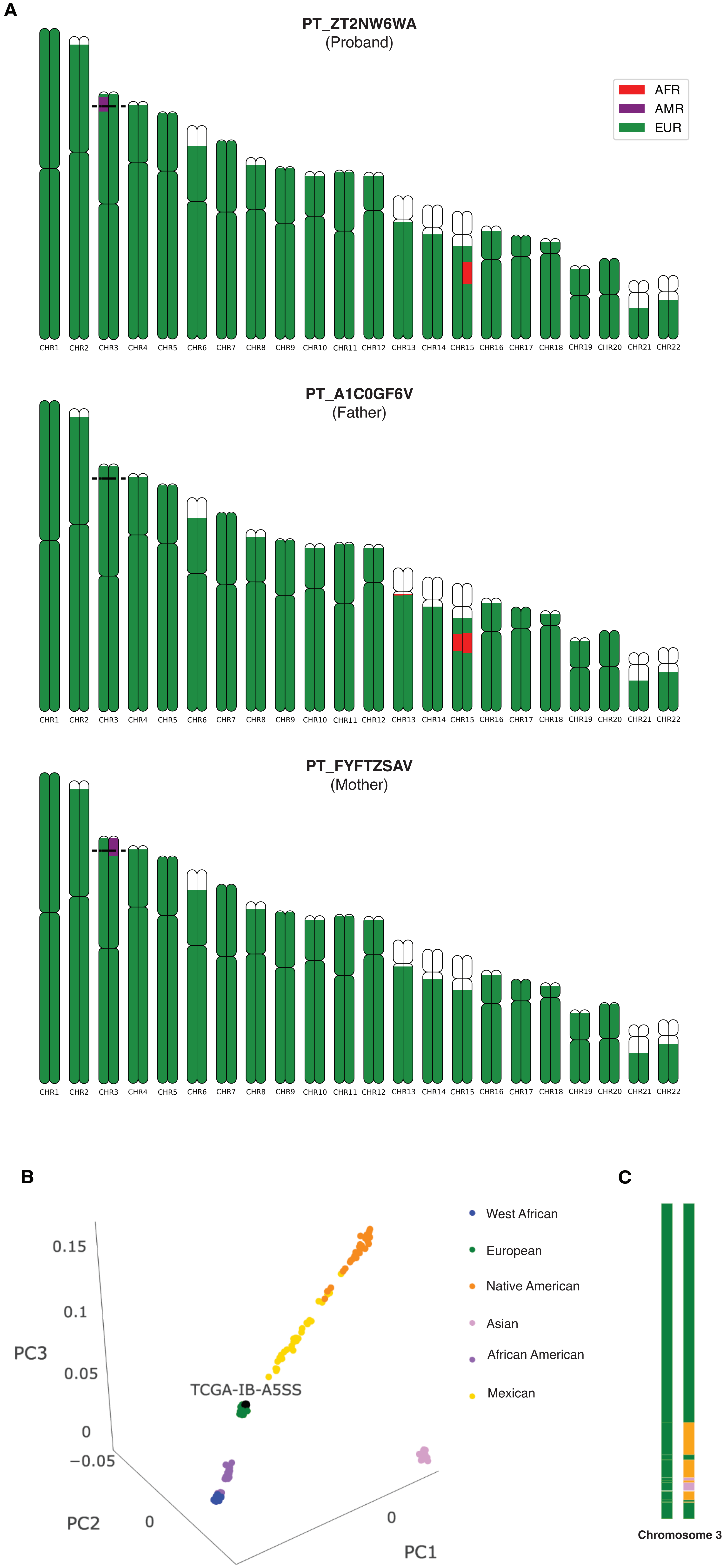

### Fig. S4

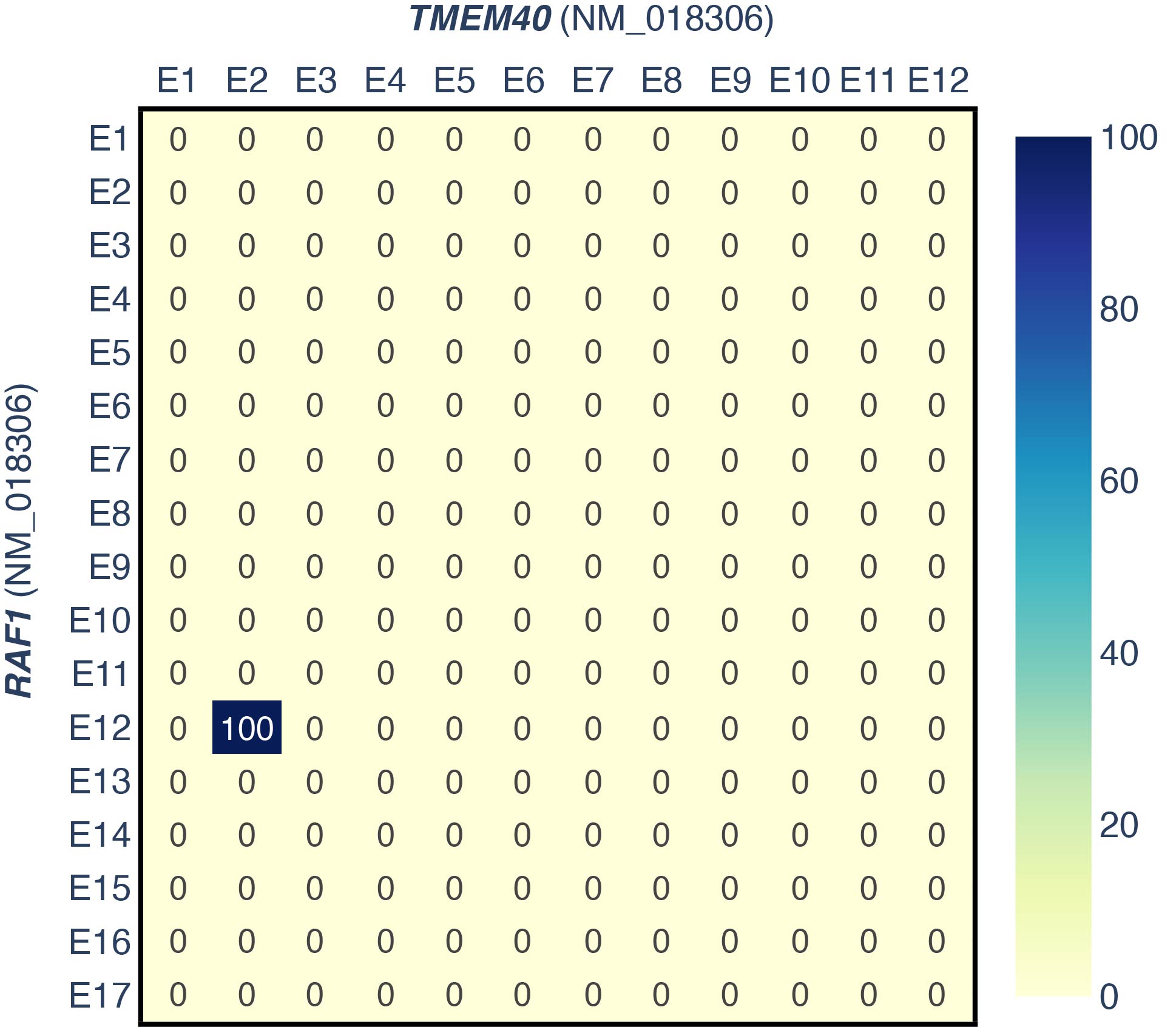

### Fig. S5

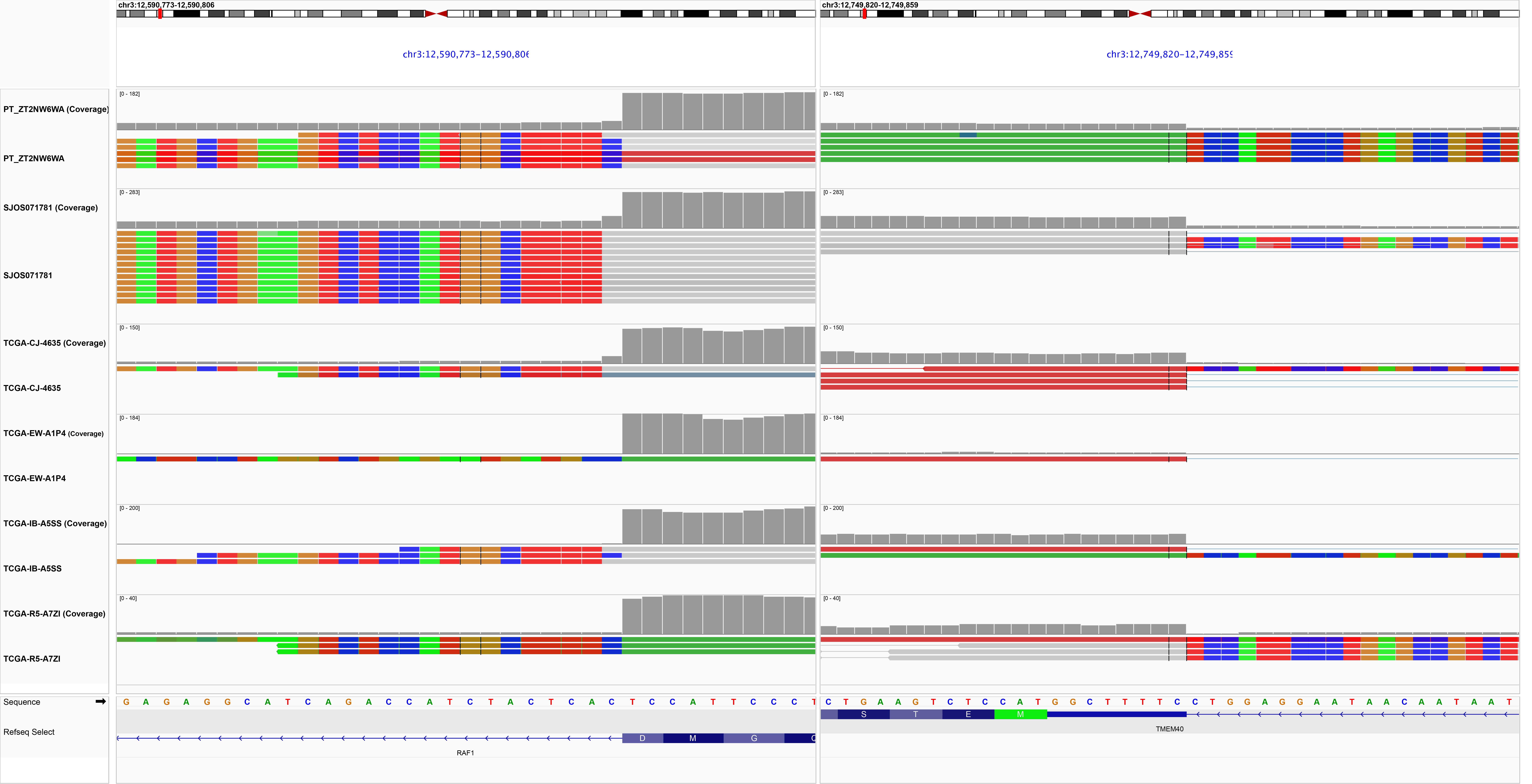
